# Descriptive epidemiology of cardiorespiratory fitness in UK adults: The Fenland Study

**DOI:** 10.1101/2022.03.01.22271683

**Authors:** Tomas I. Gonzales, Kate Westgate, Stefanie Hollidge, Tim Lindsay, Katrien Wijndaele, Nita G. Forouhi, Simon Griffin, Nick Wareham, Soren Brage

## Abstract

**Background:** Cardiorespiratory fitness is rarely measured in population studies. Most studies of fitness do not examine differences by population subgroups or seasonal trends.

**Methods:** We used a validated submaximal exercise test to measure fitness in 5976 women and 5316 men, residing in England. We expressed fitness as maximal oxygen consumption per kilogram total body mass (VO_2_max_tbm_) and fat free mass (VO_2_max_ffm_). Descriptive statistics were computed across anthropometric and sociodemographic characteristics, as well as across the year. Progressive multivariable analyses were performed to examine mediation by physical activity energy expenditure (PAEE) and BMI.

**Results:** Mean±SD VO_2_max_tbm_ was lower in women (35.4±7.6 ml·min^-1^·kg^-1^) than men (42.1±7.4 ml·min^-1^·kg^-1^) but VO_2_max_ffm_ was similar (women: 59.7±11.8 ml·min^-1^·kg^-1^; men: 62.5±10.4 ml·min^-1^·kg^-1^). Fitness was inversely associated with age but not after adjustment for PAEE. People in more physically demanding jobs were fitter compared to those in sedentary jobs but this association was attenuated in women and reversed in men following adjustment for total PAEE. PAEE and BMI and were positively associated with fitness at all levels of adjustment when fitness was expressed relative to fat-free mass. Fitness during summer was 4% higher than the winter among women, but did not differ by season among men.

**Conclusions:** Fitness was inversely associated with age but less steeply than anticipated, suggesting older generations are comparatively fitter than younger generations. PAEE and BMI were stronger determinants of the variance in fitness than any other characteristic including age. This emphasizes the importance of modifiable physical activity behaviours in public health interventions.

**KEY MESSAGES:**

- Fitness was inversely associated with age but less steeply than anticipated, suggesting older generations are comparatively fitter than younger generations
- Relationships between cardiorespiratory fitness and sociodemographic characteristics were primarily mediated by physical activity.
- A one standard deviation difference in physical activity had the same impact on cardiorespiratory fitness as being 25 years younger

## INTRODUCTION

Cardiorespiratory fitness (referred to hereafter as ‘fitness’) is inversely related to mortality and cardiometabolic disease risk ^1–5^ but is not widely recognised as a clinical vital sign in the UK. Most UK primary care providers do not routinely measure fitness, and only a few epidemiological studies have documented fitness levels in UK population subgroups. The Welsh Heart Health Survey ^6^ and Tuxworth et al ^7^ are the earliest epidemiological studies of fitness in UK adults. The Allied Dunbar National Fitness Survey was the first to establish normative fitness data for the UK population ^8^. These data were extended by the Northern Ireland Health and Activity Survey ^9^ and the 2008 Health Survey for England ^10^. The above studies have several strengths: they use dynamic exercise testing to measure differences in fitness levels by anthropometric characteristics. Exercise test selection bias limits generalisability of their findings to the UK population, however, and data on relationships between fitness, sociodemographic characteristics, and clinical characteristics are scarce. It is also unclear how these relationships may be mediated through modifiable behaviours, such as physical activity. These limitations impede public health action for improved fitness in the population. Here we examine how fitness levels vary by anthropometric, sociodemographic, and behavioural characteristics in a population-based cohort of UK adults (the Fenland Study).

## METHODS

### Study population

The Fenland Study included 12 435 participants born between 1950 and 1975 and recruited from general practice lists around Cambridgeshire, UK from January 2005 until April 2015, as described in more detail elsewhere ^11^. Exclusion criteria for participation in the Fenland study were prevalent diabetes, pregnancy or lactation, inability to walk unaided for at least 10 minutes, psychosis, or terminal illness. Participant eligibility for exercise testing was assessed using a 12-lead resting ECG (Seca CT6i), excluding those presenting with unstable angina. The present analysis included 5976 women and 5316 men with available data on fitness. The Health Research Authority NRES Committee East of England-Cambridge Central approved the study in accordance with the Declaration of Helsinki. All participants gave written informed consent. The Fenland Study has a dedicated Patient and Public Involvement panel, who provided input on the acceptability of the study protocols and participant data confidentiality. This study complied with the items listed in the Strengthening the Reporting of Observational Studies in Epidemiology (STROBE) guidelines.

### Anthropometric, sociodemographic, clinical characteristics, and physical activity

During the Fenland clinic visit, height was measured with a rigid stadiometer (SECA 240; Seca, Birmingham, UK). Total body mass was measured in light clothing with calibrated scales (TANITA model BC-418 MA; Tanita, Tokyo, Japan). Fat-free mass was measured using dual-energy x-ray absorptiometry (DEXA; Lunar-DPX) ^12^. Self-report questionnaires were used to determine sociodemographic characteristics, including participant sex, age, ethnicity (White, South Asian, Black, East Asian, other or unknown), education level (basic – compulsory schooling, further - A level/apprenticeship/sub-degree level, higher - degree level or above), work type (sedentary, standing, manual, retired, unemployed, unknown), annual household income level (<£20 000, £20 000 - £40 000, >£40 000), marital status (single, married/living as married, widowed/separated/divorced), smoking status (never, ex-smoker, current), and testing location (Cambridge, Ely, Wisbech). Resting heart rate was measured while supine using a 12-lead ECG (Seca CT6i). Following the clinic visit, objective physical activity was assessed using a combined heart rate and uniaxial movement sensor (Actiheart, CamNtech, Cambridge, UK), worn continuously for at least 72 hours and at most 6 days. Heart rate was individually calibrated ^13^ and total physical activity energy expenditure (PAEE) was computed for the wear period as described and validated elsewhere ^11,14^.

### Cardiorespiratory fitness assessment

We used an incremental, multistage, and submaximal treadmill test to estimate fitness as maximal oxygen consumption (VO_2_max). A diagram and description of the testing procedure is provided in Supplemental Figure 1. Heart rate was monitored and recorded during testing using the combined heart rate and movement sensor (Actiheart, CamNtech, Cambridge, UK) ^15^. The test ended if one of the following criteria were satisfied: 1) levelling-off of heart rate (<3bpm per min) despite an increase in work rate; 2) reaching 90% of the participant’s age-predicated maximal heart rate ^16^; 3) exercising above 80% of age-predicted max heart rate for over 2 minutes; 4) heart rate reaches 90% of the age-predicted maximal value 5) a respiratory exchange ratio of 1.1 is reached; 6) the participant wanted to stop; 7) participant indication of angina, light-headedness, or nausea; or 8) failure of the testing equipment. For participants on beta blockers, the test was terminated after 5 minutes.

To estimate VO_2_max per kg total body mass (VO_2_max_tbm_) from exercise test performance, we extrapolated the linear relationship between heart rate and work rate ^17^ to age-predicted maximal heart rate ^16^, converted the extrapolated work rate value to net VO_2_ using a caloric equivalent for oxygen ^18^, and then added an estimate of resting energy expenditure ^19^. A substudy was conducted to validate this approach against directly measured VO_2_max, demonstrating acceptable agreement (see Supplemental Methods and Results). VO_2_max per kg fat-free mass (VO_2_max_ffm_) was estimated by multiplying estimated VO_2_max_tbm_ values by total body mass and dividing by fat-free mass.

### Statistical analyses

Descriptive statistics were computed across BMI groups and sociodemographic characteristics by sex and age stratified groups. Cuzick’s test ^20^ was performed to test for trend across participant characteristics. Differences in fitness by age, BMI groups (<25, 25 to 30, and >30 kg·m^-2^) and PAEE groups (<40, 40 to 60, and >60 kJ·day^-1^ ·kg^-1^) were visualised using boxplots. Univariate associations of fitness with age, BMI, and PAEE were computed as Pearson’s r; bivariate relationships were investigated using linear regression.

We used sex-stratified and sequentially-adjusted multivariable linear regression to evaluate associations between fitness and sociodemographic characteristics (Model 1) with additional adjustment for PAEE (Model 2) and BMI (Model 3). The season of the year when fitness was measured was considered in these analyses by including two orthogonal sine functions in the regression model: “Winter” peaking at 1 on January 1^st^ and reaching a minimum of -1 on July 1^st^, and “Spring” peaking at 1 on April 1^st^ and reaching a minimum of -1 on October 1^st^. Seasonal trends were further described on a monthly basis using a binned regression procedure, controlling for seasonal variation in the measurement of sociodemographic characteristics ^21^. All analyses were performed in Stata/SE 16.1 (StataCorp, Texas, USA). Statistical significance was set at p < 0.05.

## RESULTS

In women, mean ± SD VO_2_max_tbm_ was 35.4 ± 7.6 ml O_2_·min^-1^·kg^-1^ and VO_2_max_ffm_ was 59.7 ± 11.8 ml O_2_·min^-1^·kg^-1^. In men, VO_2_max_tbm_ was 42.1 ± 7.4 ml O_2_·min^-1^·kg^-1^ and VO_2_max_ffm_ was 62.5 ± 10.4 ml O_2_·min^-1^·kg^-1^. Per five years, VO_2_max_tbm_ was lower on average by 0.2 ml O_2_·min^-1^·kg^-1^ in women and by 0.3 ml O_2_·kg^-1^·min^-1^ in men. VO_2_max_ffm_ was not associated with age in women, however in men VO_2_max_ffm_ was on average 0.05 ml O_2_·min^-1^·kg^-1^ lower per five years. Trends for other characteristics are reported in Table 1.

**Table 1.** Participant characteristics by sex-specific age groups. The Fenland Study 2005 to 2015.

| Sex | Women |  |  |  |  |  |  |  |  |
| --- | --- | --- | --- | --- | --- | --- | --- | --- | --- |
| Age group (y) | Pooled | 29-34 | 35-39 | 40-44 | 45-49 | 50-54 | 55-59 | 60-64 | P-value |
| N | 5976 | 184 | 736 | 1210 | 1322 | 1269 | 972 | 283 |  |
| Height (cm) | 164 ± 6 | 165 ± 6 | 165 ± 7 | 165 ± 6 | 165 ± 6 | 164 ± 6 | 163 ± 6 | 163 ± 6 | < 0.01 |
| Total body mass (kg) | 70.6 ± 13.9 | 67.0 ± 12.3 | 70.0 ± 15.2 | 71.0 ± 14.7 | 71.0 ± 13.8 | 70.8 ± 13.3 | 70.4 ± 13.1 | 69.7 ± 12.8 | < 0.01 |
| Body mass index (kg·m<sup>-2</sup>) | 26.2 ± 5.0 | 24.6 ± 4.2 | 25.7 ± 5.3 | 26.2 ± 5.2 | 26.2 ± 5.0 | 26.4 ± 4.8 | 26.4 ± 4.7 | 26.3 ± 4.6 | < 0.01 |
| Fat-free mass (kg) | 41.4 ± 5.2 | 40.9 ± 4.5 | 42.0 ± 5.5 | 42.1 ± 5.5 | 41.9 ± 5.2 | 41.0 ± 5.0 | 40.3 ± 5.0 | 39.9 ± 4.9 | < 0.01 |
| Resting heart rate (bpm) | 64 ± 8 | 64 ± 8 | 64 ± 9 | 64 ± 8.9 | 65 ± 9 | 64 ± 8 | 64 ± 8 | 63 ± 8 | 0.07 |
| VO<sub>2</sub>max per kg bodyweight (ml O<sub>2</sub>·min<sup>-1</sup>·kg<sup>-1</sup>) | 35.4 ± 7.6 | 36.1 ± 6.2 | 36.2 ± 7.1 | 35.8 ± 7.2 | 35.5 ± 7.7 | 35.1 ± 7.8 | 34.9 ± 8.1 | 34.7 ± 8.4 | < 0.01 |
| VO<sub>2</sub>max per kg fat-free mass (ml O<sub>2</sub>·min<sup>-1</sup>·kg<sup>-1</sup>) | 59.7 ± 11.8 | 58.5 ± 8.8 | 59.2 ± 10.2 | 59.3 ± 10.5 | 59.5 ± 11.7 | 60.0 ± 12.6 | 60.3 ± 13.3 | 60.0 ± 14.0 | 0.13 |
| PAEE (kJ·day<sup>-1</sup>·kg<sup>-1</sup>) | 51 ± 20 | 59 ± 21 | 57 ± 21 | 53 ± 20 | 51 ± 20 | 48 ± 19 | 46 ± 19 | 43 ± 16 | < 0.01 |
| Sex | Men |  |  |  |  |  |  |  |  |
| Age group (y) | Pooled | 29-34 | 35-39 | 40-44 | 45-49 | 50-54 | 55-59 | 60-64 | P-value |
| N | 5316 | 161 | 735 | 1010 | 1135 | 1072 | 929 | 274 |  |
| Height (cm) | 178 ± 7 | 179 ± 7 | 178 ± 7 | 179 ± 7 | 178 ± 7 | 177 ± 7 | 176 ± 7 | 177 ± 7 | < 0.01 |
| Bodyweight (kg) | 85.9 ± 13.9 | 84.6 ± 13.6 | 84.7 ± 14.6 | 86.2 ± 13.9 | 86.1 ± 13.9 | 86.4 ± 13.3 | 85.8 ± 13.9 | 85.4 ± 14.2 | 0.03 |
| Body mass index (kg·m<sup>-2</sup>) | 27.2 ± 4.0 | 26.4 ± 3.8 | 26.7 ± 4.2 | 27.0 ± 3.9 | 27.2 ± 4.0 | 27.5 ± 4.0 | 27.5 ± 4.0 | 27.3 ± 3.9 | < 0.01 |
| Fat-free mass (kg) | 57.3 ± 6.8 | 57.6 ± 6.8 | 57.4 ± 7.0 | 58.1 ± 6.7 | 57.6 ± 6.7 | 57.3 ± 6.7 | 56.3 ± 6.7 | 56.3 ± 7.0 | < 0.01 |
| Resting heart rate (bpm) | 61 ± 9 | 62 ± 9 | 60 ± 9 | 61 ± 9 | 61 ± 9 | 61 ± 9 | 62 ± 9 | 62 ± 9 | < 0.01 |
| VO<sub>2</sub>max per kg bodyweight (ml O<sub>2</sub>·min<sup>-1</sup>·kg<sup>-1</sup>) | 42.1 ± 7.4 | 42.5 ± 6.1 | 43.3 ± 7.0 | 43.0 ± 7.0 | 42.4 ± 7.3 | 41.3 ± 7.1 | 41.2 ± 8.2 | 40.6 ± 9.0 | < 0.01 |
| VO<sub>2</sub>max per kg fat-free mass (ml O<sub>2</sub>·min<sup>-1</sup>·kg<sup>-1</sup>) | 62.5 ± 10.4 | 61.5 ± 8.5 | 63.1 ± 9.7 | 63.2 ± 9.5 | 62.9 ± 10.4 | 62.0 ± 10.2 | 62.1 ± 11.5 | 61.2 ± 13.2 | < 0.01 |
| PAEE (kJ·day<sup>-1</sup>·kg<sup>-1</sup>) | 59 ± 23 | 65 ± 25 | 66 ± 24 | 64 ± 24 | 60 ± 23 | 57 ± 22 | 52 ± 21 | 47 ± 21 | < 0.01 |

Values are mean ± standard deviation, unless otherwise indicated. VO<sub>2</sub>max: Maximal oxygen consumption. PAEE: Physical activity energy expenditure. P-value computed from Cuzick's test for trend.

Figure 1 shows differences in VO_2_max_tbm_ and VO_2_max_ffm_ by age, BMI groups, and PAEE groups. The association of fitness with age, BMI, and PAEE was strongest when expressed as VO_2_max_tbm_ compared to VO_2_max_ffm_. We investigated this further by conducting univariate and bivariate analyses of VO_2_max_tbm_ and VO_2_max_ffm_ with BMI, PAEE, and age (Supplemental Figures 2 and 3). The association of VO_2_max_tbm_ with PAEE (Pearson’s r for women: 0.39; men: r: 0.39) was higher than associations with BMI (Pearson’s r for women: - 0.34; men: -0.23) and age (Pearson’s r for women: -0.06; men: -0.11). The combination of BMI and PAEE explained more variance in VO_2_max_tbm_ (20% for women, 18% for men) than bivariate combinations with age. For VO_2_max_ffm_, univariate and bivariate analyses had weaker associations and less explained variance than analogous results for VO_2_max_tbm_.

**Figure 1.**
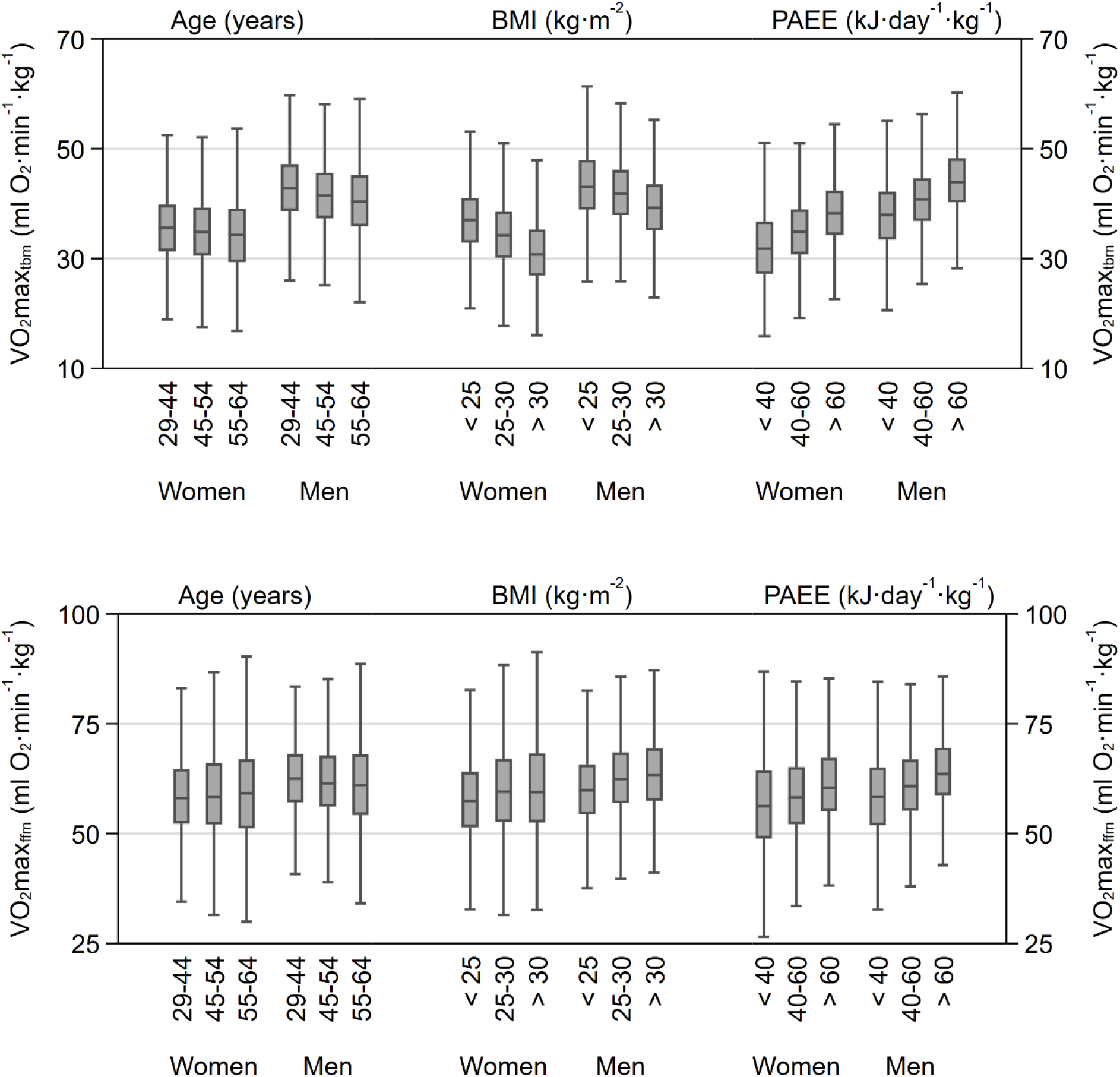
Sex stratified maximal oxygen consumption per kilogram total body mass (VO_2_max_tbm_; top panel) and per kilogram fat free mass (VO_2_max_ffm_; bottom panel) by age, body mass index (BMI), and physical activity energy expenditure (PAEE). Box plots represent medians, interquartile ranges, and minimum– maximum ranges without outliers. The Fenland Study 2005 to 2015.

Unadjusted mean VO_2_max_tbm_ values, stratified by sex and sociodemographic characteristics, are provided in Table 2. VO_2_max_tbm_ was generally higher in women with more educational attainment (Higher education: 36.2 ± 7.3 ml O_2_·min^-1^·kg^-1^; No education: 34.6 ± 8.6 ml O_2_·min^-1^·kg^-1^), but did not differ by education in men (Higher education: 42.1 ± 7.2 ml O_2_·min^-1^·kg^-1^; No education: 42.4 ± 6.9 ml O_2_·min^-1^·kg^-1^). Workers in more physically demanding jobs (Female manual workers: 36.4 ± 8.3 ml O_2_·min^-1^·kg^-1^; male manual workers: 42.7 ± 7.9 ml O_2_·min^-1^·kg^-1^) were fitter than those in sedentary jobs (Female sedentary workers: 35.1 ± 7.2 ml O_2_·min^-1^·kg^-1^; male sedentary workers: 41.8 ± 7.0 ml O_2_·min^-1^·kg^-1^). Current smokers had higher VO_2_max_tbm_ (Female smokers: 38.1 ± 8.5 ml O_2_·min^-1^·kg^-1^; male smokers: 44.2 ± 8.1 ml O_2_·min^-1^·kg^-1^) than non-smokers (Female non-smokers: 35.0 ± 7.4 ml O_2_·min^-1^·kg^-1^; male non-smokers: 41.8 ± 7.2 ml O_2_·min^-1^·kg^-1^). VO_2_max_tbm_ differed by ethnicity, however sample sizes were disproportionate between whites and other racial and ethnic groups (See Supplemental Tables 1 and 2). VO_2_max_tbm_ differences were not apparent between income levels, marital status, and testing sites. Analogous results for VO_2_max_ffm_ are presented in Table 3.

**Table 2.**
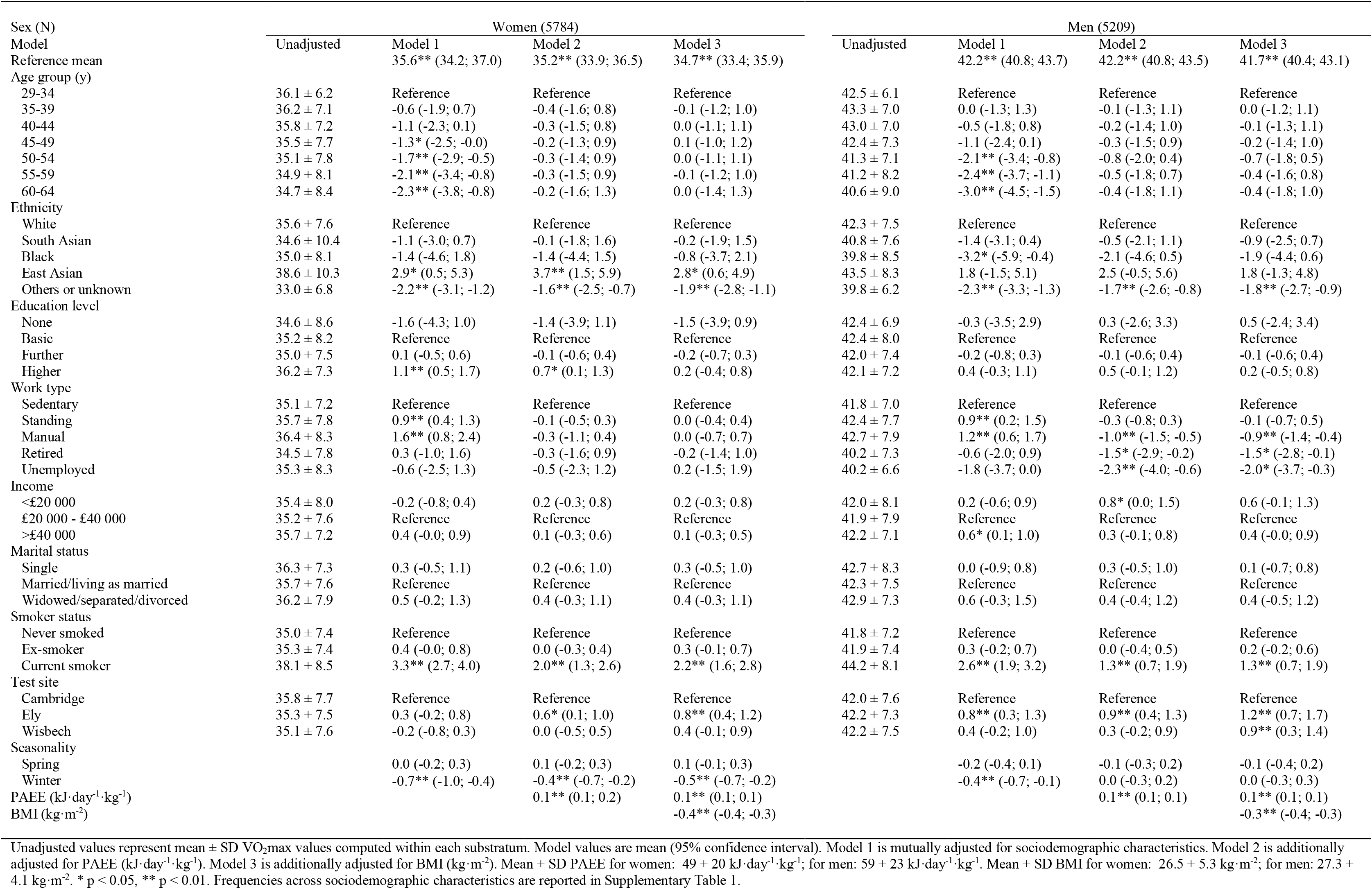
Sequentially-adjusted multivariable analysis of maximal oxygen consumption (VO_2_max) per kg total body mass (ml O_2_·min^-1^·kg^-1^) by sociodemographic characteristics. The Fenland Study 2005 to 2015.

**Table 3.** Sequentially-adjusted multivariable analysis of maximal oxygen consumption (VO_2_max) per kg fat free mass (ml O_2_·min^-1^·kg^-1^) by sociodemographic characteristics. The Fenland Study 2005 to 2015.

| Sex (N) | Women (5530) |  |  |  | Men (4980) |  |  |  |
| --- | --- | --- | --- | --- | --- | --- | --- | --- |
|  | Unadjusted | Model 1 | Model 2 | Model 3 | Unadjusted | Model 1 | Model 2 | Model 3 |
| Reference mean |  | 59.3** (57.0; 61.5) | 59.0** (56.8; 61.2) | 59.5** (57.3; 61.7) |  | 61.5** (59.4; 63.7) | 61.5** (59.4; 63.6) | 62.2** (60.1; 64.3) |
| Age group (y) |  |  |  |  |  |  |  |  |
| 29-34 | 58.5 ± 8.8 | Reference | Reference | Reference | 61.5 ± 8.5 | Reference | Reference | Reference |
| 35-39 | 59.2 ± 10.2 | -0.7 (-2.8; 1.3) | -0.6 (-2.6; 1.4) | -0.9 (-2.9; 1.1) | 63.1 ± 9.7 | 0.4 (-1.6; 2.3) | 0.3 (-1.6; 2.1) | 0.1 (-1.7; 2.0) |
| 40-44 | 59.3 ± 10.5 | -0.9 (-2.9; 1.1) | -0.3 (-2.3; 1.6) | -0.7 (-2.6; 1.3) | 63.2 ± 9.5 | 0.2 (-1.7; 2.1) | 0.4 (-1.5; 2.3) | 0.2 (-1.6; 2.1) |
| 45-49 | 59.5 ± 11.7 | -0.6 (-2.6; 1.3) | 0.1 (-1.9; 2.0) | -0.2 (-2.1; 1.7) | 62.9 ± 10.4 | -0.1 (-2.0; 1.8) | 0.5 (-1.3; 2.4) | 0.4 (-1.5; 2.2) |
| 50-54 | 60.0 ± 12.6 | -0.1 (-2.1; 1.9) | 0.9 (-1.1; 2.8) | 0.6 (-1.4; 2.5) | 62.0 ± 10.2 | -1.1 (-3.0; 0.8) | -0.0 (-1.9; 1.9) | -0.2 (-2.1; 1.6) |
| 55-59 | 60.3 ± 13.3 | -0.2 (-2.3; 1.8) | 1.0 (-1.0; 3.1) | 0.8 (-1.2; 2.9) | 62.1 ± 11.5 | -1.2 (-3.2; 0.7) | 0.3 (-1.6; 2.2) | 0.1 (-1.7; 2.0) |
| 60-64 | 60.0 ± 14.0 | -0.6 (-3.1; 1.8) | 0.9 (-1.5; 3.3) | 0.8 (-1.6; 3.2) | 61.2 ± 13.2 | -2.0 (-4.2; 0.3) | 0.2 (-1.9; 2.4) | 0.2 (-2.0; 2.4) |
| Ethnicity |  |  |  |  |  |  |  |  |
| White | 59.9 ± 11.8 | Reference | Reference | Reference | 62.7 ± 10.5 | Reference | Reference | Reference |
| South Asian | 60.2 ± 15.4 | 1.3 (-1.6; 4.3) | 2.0 (-0.9; 5.0) | 2.1 (-0.8; 4.9) | 62.7 ± 12.0 | 0.3 (-2.2; 2.7) | 1.0 (-1.4; 3.4) | 1.5 (-0.8; 3.9) |
| Black | 56.7 ± 13.1 | -2.9 (-7.9; 2.1) | -3.0 (-7.9; 2.0) | -3.6 (-8.5; 1.3) | 55.3 ± 12.1 | -7.6** (-11.6; -3.6) | -6.9** (-10.8; -3.0) | -7.2** (-11.0; -3.4) |
| East Asian | 61.7 ± 15.5 | 3.0 (-0.8; 6.7) | 3.5 (-0.2; 7.2) | 4.4* (0.7; 8.0) | 62.3 ± 12.1 | 1.2 (-3.4; 5.9) | 1.8 (-2.7; 6.4) | 2.9 (-1.6; 7.4) |
| Others or unknown | 55.0 ± 10.3 | -3.6** (-5.2; -2.0) | -3.3** (-4.9; -1.7) | -3.0** (-4.6; -1.5) | 59.4 ± 8.3 | -2.4** (-4.0; -0.9) | -1.9* (-3.4; -0.4) | -1.9* (-3.4; -0.4) |
| Education level |  |  |  |  |  |  |  |  |
| None | 60.0 ± 15.4 | -2.7 (-7.1; 1.7) | -2.8 (-7.2; 1.5) | -2.9 (-7.2; 1.4) | 63.5 ± 11.0 | -0.1 (-5.0; 4.7) | 0.8 (-3.9; 5.5) | 0.4 (-4.3; 5.0) |
| Basic | 60.7 ± 13.2 | Reference | Reference | Reference | 63.7 ± 10.9 | Reference | Reference | Reference |
| Further | 59.5 ± 11.8 | -0.5 (-1.3; 0.3) | -0.6 (-1.5; 0.2) | -0.5 (-1.3; 0.3) | 62.8 ± 10.6 | -0.4 (-1.3; 0.4) | -0.4 (-1.2; 0.5) | -0.4 (-1.2; 0.4) |
| Higher | 59.2 ± 10.9 | -0.5 (-1.5; 0.4) | -0.9 (-1.8; 0.1) | -0.4 (-1.3; 0.6) | 61.7 ± 10.0 | -0.8 (-1.8; 0.2) | -0.7 (-1.7; 0.3) | -0.3 (-1.3; 0.7) |
| Work type |  |  |  |  |  |  |  |  |
| Sedentary | 59.1 ± 11.1 | Reference | Reference | Reference | 62.1 ± 9.8 | Reference | Reference | Reference |
| Standing | 60.1 ± 12.2 | 0.8* (0.1; 1.5) | 0.1 (-0.7; 0.8) | 0.0 (-0.7; 0.7) | 63.3 ± 11.1 | 0.9 (-0.0; 1.8) | 0.0 (-0.9; 0.8) | -0.2 (-1.1; 0.7) |
| Manual | 61.2 ± 13.7 | 1.6* (0.4; 2.9) | 0.3 (-1.0; 1.5) | 0.0 (-1.2; 1.2) | 63.2 ± 11.0 | 0.3 (-0.5; 1.1) | -1.6** (-2.3; -0.8) | -1.7** (-2.5; -0.9) |
| Retired | 60.2 ± 13.8 | 0.7 (-1.4; 2.7) | 0.2 (-1.8; 2.2) | 0.0 (-2.0; 2.0) | 60.2 ± 10.6 | -1.6 (-3.7; 0.4) | -2.4* (-4.5; -0.4) | -2.5* (-4.5; -0.5) |
| Unemployed | 61.1 ± 13.1 | 0.7 (-2.3; 3.8) | 0.7 (-2.3; 3.7) | 0.1 (-2.9; 3.1) | 60.2 ± 9.3 | -3.0* (-5.7; -0.3) | -3.2* (-5.8; -0.6) | -3.7** (-6.3; -1.1) |
| Income |  |  |  |  |  |  |  |  |
| <£20 000 | 60.6 ± 12.8 | 0.2 (-0.8; 1.2) | 0.5 (-0.5; 1.5) | 0.6 (-0.4; 1.5) | 63.0 ± 11.9 | 0.6 (-0.5; 1.8) | 1.1* (0.0; 2.2) | 1.3* (0.2; 2.4) |
| £20 000 - £40 000 | 59.8 ± 12.0 | Reference | Reference | Reference | 62.4 ± 11.0 | Reference | Reference | Reference |
| >£40 000 | 59.2 ± 11.0 | 0.1 (-0.6; 0.9) | -0.1 (-0.8; 0.6) | 0.0 (-0.8; 0.7) | 62.5 ± 9.8 | 0.8* (0.1; 1.5) | 0.6 (-0.1; 1.3) | 0.5 (-0.2; 1.2) |
| Marital status |  |  |  |  |  |  |  |  |
| Single | 60.8 ± 11.9 | 0.7 (-0.6; 2.0) | 0.6 (-0.7; 1.9) | 0.5 (-0.7; 1.8) | 62.6 ± 12.0 | -0.3 (-1.5; 0.8) | -0.1 (-1.2; 1.0) | 0.2 (-0.9; 1.3) |
| Married/living as married | 60.0 ± 11.9 | Reference | Reference | Reference | 62.8 ± 10.5 | Reference | Reference | Reference |
| Widowed/separated/divorced | 61.1 ± 12.4 | 0.5 (-0.7; 1.6) | 0.3 (-0.8; 1.5) | 0.3 (-0.9; 1.5) | 63.3 ± 10.5 | 0.4 (-0.9; 1.7) | 0.2 (-1.1; 1.4) | 0.2 (-1.0; 1.5) |
| Smoker status |  |  |  |  |  |  |  |  |
| Never smoked | 58.9 ± 11.5 | Reference | Reference | Reference | 61.7 ± 10.0 | Reference | Reference | Reference |
| Ex-smoker | 59.7 ± 11.7 | 0.7* (0.1; 1.4) | 0.5 (-0.2; 1.2) | 0.3 (-0.4; 0.9) | 62.8 ± 10.4 | 1.2** (0.5; 1.8) | 1.0** (0.4; 1.6) | 0.7* (0.1; 1.3) |
| Current smoker | 63.7 ± 13.3 | 4.6** (3.5; 5.7) | 3.6** (2.5; 4.6) | 3.4** (2.4; 4.4) | 65.2 ± 11.7 | 3.5** (2.5; 4.4) | 2.4** (1.5; 3.3) | 2.4** (1.5; 3.3) |
| Test site |  |  |  |  |  |  |  |  |
| Cambridge | 59.5 ± 11.7 | Reference | Reference | Reference | 61.7 ± 10.6 | Reference | Reference | Reference |
| Ely | 59.1 ± 11.9 | 0.5 (-0.3; 1.3) | 0.7 (-0.0; 1.5) | 0.5 (-0.2; 1.3) | 62.4 ± 10.0 | 1.1** (0.4; 1.9) | 1.2** (0.5; 1.9) | 0.7* (0.0; 1.5) |
| Wisbech | 60.8 ± 11.9 | 1.0* (0.2; 1.9) | 1.2** (0.3; 2.1) | 0.8 (-0.0; 1.7) | 64.0 ± 10.7 | 2.1** (1.2; 2.9) | 2.0** (1.1; 2.8) | 1.2** (0.4; 2.1) |
| Seasonality |  |  |  |  |  |  |  |  |
| Spring |  | -0.1 (-0.5; 0.4) | -0.1 (-0.5; 0.4) | -0.1 (-0.5; 0.3) |  | -0.4 (-0.8; 0.0) | -0.3 (-0.7; 0.1) | -0.3 (-0.7; 0.1) |
| Winter |  | -1.4** (-1.9; -1.0) | -1.3** (-1.7; -0.8) | -1.2** (-1.6; -0.8) |  | -0.4 (-0.8; 0.0) | -0.1 (-0.5; 0.3) | -0.1 (-0.5; 0.3) |
| PAEE (kJ·day<sup>-1</sup>·kg<sup>-1</sup>) |  |  | 0.1** (0.1; 0.1) | 0.1** (0.1; 0.1) |  |  | 0.1** (0.1; 0.1) | 0.1** (0.1; 0.1) |
| BMI (kg·m<sup>-2</sup>) |  |  |  | 0.4** (0.3; 0.4) |  |  |  | 0.5** (0.4; 0.5) |

Sequentially adjusted multivariable analyses of associations between VO_2_max_tbm_ and sociodemographic characteristics are reported in Table 2. Age was still inversely associated with VO_2_max_tbm_ but was attenuated to the null with adjustment for PAEE. Occupation type was not associated with differences in VO_2_max_tbm_ among women after adjustment. Among men, VO_2_max_tbm_ was lower in manual workers (−0.9 ml O_2_·min^-1^·kg^-1^, 95% CI: -1.4 to -0.4), retirees (−1.5 ml O_2_·min^-1^·kg^-1^, 95% CI: -2.8 to -0.1), and the unemployed (−2.0 ml O_2_·min^- 1^·kg^-1^, 95% CI: -3.7 to -0.3) relative to sedentary workers. Current smokers still had higher VO_2_max_tbm_ (women: 2.2 ml O_2_·min^-1^·kg^-1^, 95% CI: 1.6 to 2.8; men: 1.3 ml O_2_·min^-1^·kg^-1^, 95% CI: -0.7 to 1.9) relative to non-smokers. VO_2_max_tbm_ did not differ by education level, income, and marital status in both women and men after adjustment. Table 3 presents analogous results for VO_2_max_ffm_. The direction and magnitude of differences in VO_2_max_ffm_ across sociodemographic factors were largely similar to those found for VO_2_max_tbm_, however Black men had lower VO_2_max_ffm_ (−7.2 ml O_2_·min^-1^·kg^-1^, 95% CI: -11.0 to -3.4) relative to White men.

Women tested in the winter had lower VO_2_max_tbm_ and VO_2_max_ffm_ than those tested at other times of the year. Seasonal variation in VO_2_max_tbm_ and VO_2_max_ffm_ was not observed in men after adjustment for PAEE. To investigate this further, we analysed seasonal variation in fitness when stratified by higher (≥50 kJ·day^-1^·kg^-1^) and lower (<50 kJ·day^-1^·kg^-1^) PAEE levels (Figure 2). Seasonal fitness variation persisted in men and women with higher PAEE levels. Women with lower PAEE levels also demonstrated seasonal variation, however fitness measurements did not differ by season in men with lower PAEE.

**Figure 2.**
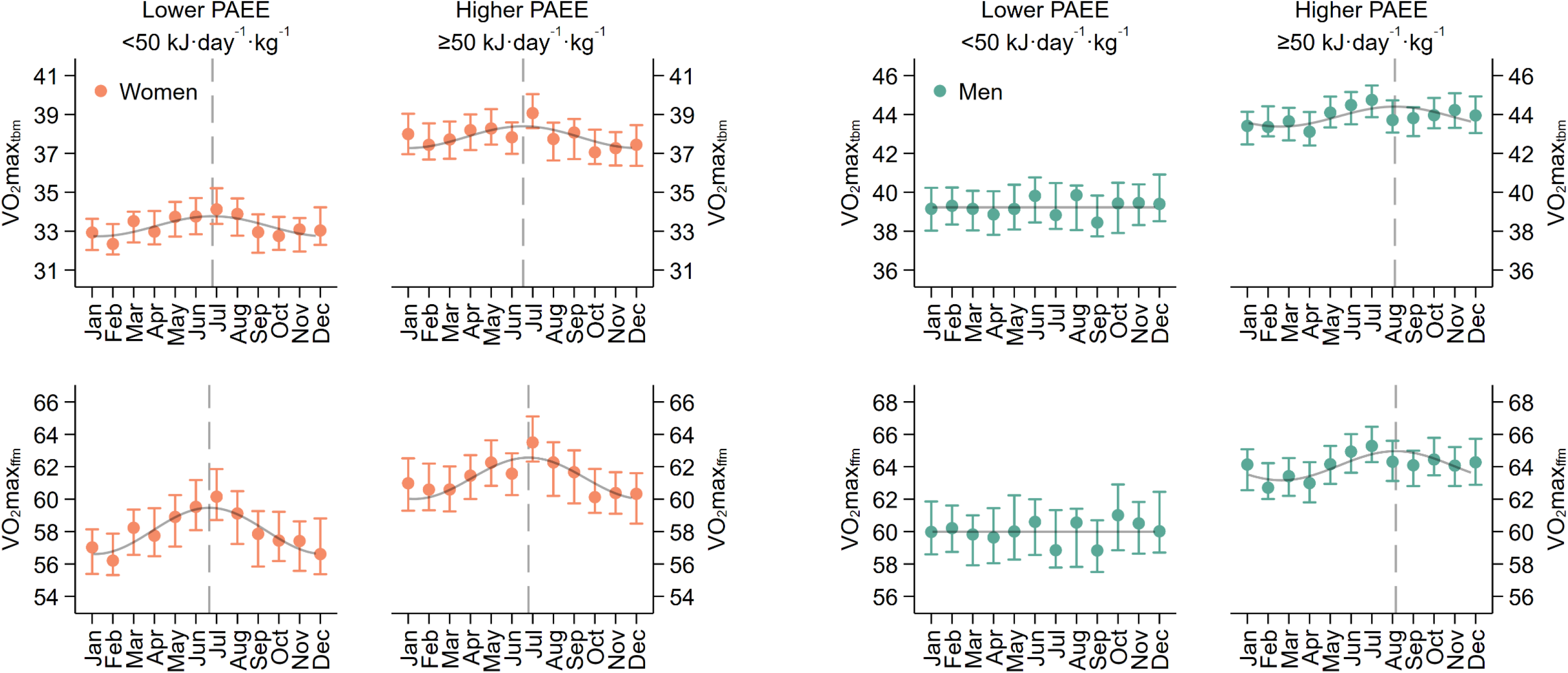
Seasonal variation in maximal oxygen consumption per kilogram total body mass (VO_2_max_tbm_) and per kilogram fat free mass (VO_2_max_ffm_) stratified by sex and physical activity energy expenditure (PAEE) levels. Dots and bars represent point estimates and 95% confidence intervals from a binned regression procedure, adjusted for age, ethnicity, education level, work type, income, marital status, smoker status, and fitness testing site. Superimposed curves represent seasonal fitness values derived from an unadjusted cosinor model. Vertical dashed lines represent seasonal peaks in fitness values where seasonal variation is observed.

## DISCUSSION

Here we have examined how fitness varies by anthropometric, sociodemographic, and behavioural characteristics in a population-based sample of UK adults (The Fenland Study). Physical activity was a more important determinant of fitness than chronological age, highlighting the importance of maintaining an active lifestyle in adulthood. Our findings extend previous reports of fitness in the UK and provide directions for future population-based studies of fitness.

Fitness was lower in older versus younger UK adults, however the magnitude of difference per five years (approximately 0.3 ml O_2_·kg^-1^·min^-1^) was less than values published in some national fitness registries. Fitness declined by approximately 2 ml O_2_·kg^-1^·min^-1^ per five years in the US-based FRIEND registry ^22^, Brazil-based Fleury study ^23^, and Norway-based HUNT study ^24^. Similarly, the German-based Prevention First Registry was 1.5 ml O_2_·kg^-1^·min^-1^, German-based SHIP study ^25^ was 1 ml O_2_·kg^-1^·min^-1^, and both the Danish Health Examination Survey ^26^ and Korean-based KISS FitS study ^27^ were 1.3 ml O_2_·kg^-1^·min^-1^. These values are similar to those reported by previous UK-based population studies of fitness ^5–9^, where absolute fitness values were generally lower than those reported in the present study. While it is possible that fitness in UK adults has improved compared to previous UK studies of fitness, the difference in the magnitude of fitness decline with age between those and the present study may also be due to participant selection bias in the Fenland cohort. Healthy and enthusiastic people may have volunteered for study participation more often than unhealthy people (the “healthy participant effect”). We investigated this in a previous study ^11^, finding the Fenland cohort to be slightly healthier than the population from which they were sampled. Thus, it is possible that higher fitness values reported here reflect regional differences in health across the UK.

The association between fitness and physical activity was stronger than that for age; a one standard deviation higher PAEE equates to the same difference in fitness (∼2 ml O_2_·kg^-1^·min^-1^ higher) as being 25 years younger. Similarly, a one standard deviation higher BMI was associated with lower fitness (1.2 – 1.6 ml O_2_·kg^-1^·min^-1^), adjusted for age, physical activity and other sociodemographic characteristics. Thus, physical activity and BMI were stronger determinants of fitness than age and other factors examined. Physical activity is known to improve and explain a majority of the variance in fitness among adults ^28–30^, but age-related decline in fitness is not wholly due to physical inactivity ^31,32^. Reduced cardiac output and impaired skeletal muscle oxidative capacity with age are also contributing factors, particularly after the seventh decade of life ^33–35^. We did not directly measure cardiac output or skeletal muscle function, and therefore cannot investigate whether the preservation of fitness with age is related to maintenance of these factors. We also did not measure fitness in adults aged 70 years and over, when the impact of higher physical activity on improved fitness may wane. Nevertheless, our data suggest that higher physical activity can alter the trajectory of fitness decline with age in generally healthy adults. It is unclear whether this finding is reflective of health promotion strategies to increase physical activity and fitness within the Cambridgeshire region ^36^. If so, future work could seek how these strategies may be adapted to UK regions with high cardiometabolic disease prevalence.

People in more physically demanding occupations were fitter than those in sedentary occupations. When accounting for PAEE and BMI, however, fitness did not differ by occupation in women; in men, fitness was statistically lower in manual, retired, and unemployed workers compared to sedentary workers. Other studies report that manual workers may have greater muscle strength but less overall fitness than the general population ^37,38^. While it is not immediately clear as to the mechanism by which manual work would lower fitness in men, it is likely occupation specific and could be related to diminished lung function ^39^. Previously we reported that manual workers in the Fenland cohort had greater physical activity levels than other occupation types ^11^. It is therefore reassuring to observe that the negative effect of manual work on fitness - whatever the mechanism – is partly ameliorated by higher physical activity among male workers. Alternatively, the observed association between low fitness and manual work could be due to residual confounding for socioeconomic status. Previous research suggest that lower fitness in retired and unemployed male workers is related to advanced age and cardiovascular deconditioning after long-lasting physical inactivity ^40^.

We present fitness results scaled by both total body mass (VO_2_max_tbm_) and fat free mass (VO_2_max_ffm_). In multivariable analysis, VO_2_max_tbm_ was negatively associated with BMI, however VO_2_max_ffm_ was positively associated. Other studies demonstrate that fitness is independent of adiposity when scaled by fat free mass and suggest VO_2_max_ffm_ can be considered an indirect measure of musculoskeletal tissue metabolic quality ^41,42^. More direct measurements of muscle oxidative capacity, such as tissue biopsy or imaging ^43^, could be used to elucidate whether this is preserved in otherwise overweight and obese participants with higher VO_2_max_ffm_ values. Indeed, ectopic fat infiltration of skeletal muscle may be more related to impaired muscle oxidative capacity and reduced force production than overall adiposity ^44,45^.

Fitness measurements were generally higher in the summer compared to the spring and winter. Physical activity measurements had a similar pattern in previous analyses ^11^. PAEE adjustment negated seasonal variation of fitness in men, however in women seasonal variation persisted. Given the cross-sectional analysis used in this study, we recognise that the results regarding fitness seasonality should be interpreted cautiously and a repeated measures design could be more appropriate. Such a design would be largely unfeasible for a population-based study of fitness, however, since increased test frequency would be costly to scale and would increase lost-to-follow-up rates. Future work could elucidate whether seasonal variation in fitness among women is related to seasonal variation in endogenous factors such as circannual hormonal rhythms ^46^.

Our study has strengths and limitations. We objectively assessed fitness in a large participant sample, enabling the investigation of differences by sociodemographic characteristics in the UK. We also quantified and compared the influence of objectively measured PAEE and BMI on associations between fitness and these characteristics, which allow judgement of their relative importance. A limitation of our study includes using heart rate response to a submaximal exercise test, rather than directly measured maximal oxygen consumption; however, as we show here, the fitness estimates from this method agree with direct measurements which provides reassurance of our findings. Another potential limitation is the non-representativeness of the Fenland cohort compared to the random population sampling frame. Compared to non-responders, participants were slightly older, leaner, less likely to smoke, more likely to drink alcohol, and more likely to live in deprived neighbourhoods; these differences were small, however. In addition, observed physical activity levels - the strongest determinant of fitness - were similar to those observed in the general UK population ^11,47^, suggesting findings may generalise more widely.

We have described variation in fitness within a UK adult population by sociodemographic factors and lifestyle behaviours. Fitness was inversely associated with age but less steeply than anticipated, suggesting older generations are comparatively fitter than younger generations. Physical activity and body size were stronger determinants of the variance in fitness than any other factor including age. A one standard deviation difference in physical activity had the same impact on cardiorespiratory fitness as being 25 years younger. This emphasizes the importance of maintaining physical activity across adulthood.

## Data Availability

The datasets generated and analysed during the current study are available at request via the MRC Epidemiology website.

http://www.mrc-epid.cam.ac.uk/research/data-sharing/

## ETHICS APPROVAL

The participants in the Fenland study were recruited from general practice lists as the population-based sampling frame. The National Research Ethics Service (NRES), the body that approves the ethics of research involving NHS patients, considered and approved the study through its East of England Cambridge Central Committee in accordance with the Declaration of Helsinki. All participants provided written informed consent.

## AUTHOR CONTRIBUTIONS

The authors contributed to the present manuscript as follows: Idea for analysis (T.G., S.B.); acquisition and analysis of cardiorespiratory fitness data (K.We., S.H., S.B.); acquisition and analysis of raw physical activity data (K.We., S.H., S.B.); analysis of epidemiological data (T.G., T.L.); drafting of the manuscript (T.G.); revising work critically for important intellectual content (all authors); approval of the final version before submission (all authors). Chief Investigator (N.W.) and Principal Investigators (NF, SG, SB) of the Fenland Study.

## DATA AVAILABILITY

The datasets generated and analysed during the current study are available at request via the MRC Epidemiology website (http://www.mrc-epid.cam.ac.uk/research/data-sharing/).

## SUPPLEMENTARY DATA

Supplementary data are available at IJE online.

## FUNDING

The Fenland study was funded by the Medical Research Council and the Wellcome Trust. The current work was supported by the Medical Research Council (T.G., S.B., K.Wi., S.H., grant number MC_UU_12015/3), (S.G., grant number MC_UU_12015/4), (N.W., grant number MC_UU_12015/1), (N.G.F., grant number MC_UU_12015/5); the National Institute of Health Research Cambridge (NIHR) Biomedical Research Centre (K.We., S.B., N.G.F., and N.W., grant number IS-BRC-1215-20014); and the Cambridge Trust and St Catharine’s College (T.L.). The funders had no role in the design, analysis or writing of this manuscript.

## ACKNOWLEDGEMENTS

We are grateful to the Fenland Study participants for their willingness and time to take part. We thank all members of the following teams responsible for practical aspects of the study; Study Coordination, Field Epidemiology, Anthropometry Team, Physical Activity Technical Team, IT, Data Management, and Statistics.

## CONFLICT OF INTEREST

None declared.

## Supplemental Materials

### Validation sub-study participant characteristics

We examined the validity of several methods for predicting VO_2_max from HR response to the Cambridge Ramped Treadmill Test (CRTT), a submaximal exercise test used in the present study to estimate fitness. The test was originally designed for individual calibration of HR-to-energy expenditure for the quantification of physical activity energy expenditure during free-living, but has been adapted to estimate fitness in population-based studies.

We recruited 97 participants (51 females, 46 males) for the validation study. Nine participants were excluded from the present analyses for terminating the treadmill test early and 5 participants for equipment failure, resulting in a final sample of 42 females and 41 males with valid measures of VO2max. Participant characteristics are described below:

| Characteristic | Female (n = 42) | Male (n = 41) |
| --- | --- | --- |
| Age (y) | 39.6 ± 12.3 | 41.7 ± 13.5 |
| Height (cm) | 165.1 ± 6.4 | 177.5 ± 6.9 |
| Weight (kg) | 63.4 ± 7.5 | 78.9 ± 11.7 |
| BMI (kg·m<sup>-2</sup>) | 23.2 ± 2.6 | 25.1 ± 3.6 |
| Fat free mass (kg) | 44.5 ± 3.4 | 62.9 ± 6.4 |
| RHR (bpm) | 60.9 ± 8.5 | 60.1 ± 10.7 |
| Measured HRmax (bpm) | 186.3 ± 11.4 | 186.7 ± 10.6 |
| Age-predicted HRmax (bpm) | 180.3 ± 8.6 | 178.8 ± 9.4 |
| Measured REE (J·kg<sup>-1</sup>·min<sup>-1</sup>) | 59.4 ± 7.7 | 60.6 ± 8.4 |
| Predicted REE (J·kg<sup>-1</sup>·min<sup>-1</sup>) | 62.6 ± 4.4 | 64.4 ± 4.4 |
| VO<sub>2</sub>max per kg bodyweight (ml O<sub>2</sub>·kg<sup>-1</sup>·min<sup>-1</sup>) | 39.7 ± 6.7 | 46.0 ± 9.5 |
| VO<sub>2</sub>max per kg fat-free mass (ml O<sub>2</sub>·kg<sup>-1</sup>·min<sup>-1</sup>) | 56.0 ± 7.9 | 56.8 ± 8.7 |

BMI: body mass index; RHR: resting heart rate; HRmax: maximal heart rate; REE: Resting energy expenditure; VO<sub>2</sub>max: maximal oxygen consumption; Age-predicted HRmax = 208 - 0.7 · age(y); Predicted REE was computed according to Henry (2005)

### Measurement of VO_2_, resting energy expenditure, and VO_2_max

VO_2_ during rest and during the CRTT was measured using a computerised metabolic cart (Oxycon Pro, Erich Jaeger GmbH, Hoechberg, Germany); this system has been validated previously <u>(15).</u> Energy expenditure was calculated by indirect calorimetry according to Weir <u>(16).</u> REE was measured in the supine position with a ventilated hood positioned over the participant’s head for 15 minutes; the mean of the last 5 min was used in analysis.

The CRTT was conducted according to methods described in the main study, however in the validation study the test was extended with an additional stage to allow direct measurement of VO_2_max. After Stage IV of the original protocol, treadmill speed was increased by 0.25 km·h^-1^ and incline by 0.5% every 15s until exhaustion was reached. Breath-by-breath values of VO_2_ were averaged in 15s epochs, filtering out the highest and lowest breath values, and VO_2_max was computed as the average of the two largest VO_2_ values in the last 45s of the test. The test was terminated if one of the following three criteria were satisfied: 1) the participant wanted to stop despite verbal encouragement; 2) participant indication of angina, light-headedness, or nausea; and 3) failure of the testing equipment. In analysis, VO_2_max was considered reached if two of the four following criteria were achieved: 1) respiratory exchange ratio value > 1.2; 2) leveling-off in VO_2_ (< 2.5 ml O_2_·kg^-1^·min^-1^ change) despite an increase in work rate; 3) leveling-off in HR (< 3bpm per min) despite an increase in WR; and 4) reaching 100% of the participant’s age-predicted HRmax.

### Statistical analyses

Correlations between predicted and directly measured VO_2_max were quantified using Pearson’s *r* and Spearman’s *rho*. Bias was computed as the difference between predicted and directly measured VO_2_max. One-sample t-tests were performed to determine whether mean biases were statistically significantly different from zero. Differential bias was examined by sex and across combinations of test endpoints (CRTT stages and percentages of age-predicted HRmax). Prediction precision was expressed as the root mean square error (RMSE). Statistical significance was set to 0.05.

### Results

We examined the validity of estimated VO_2_max values computed from data terminated after stages II, III, and IV, and at percentages of age-predicted HRmax. The table below demonstrates levels of agreement between predicted and directly measured VO_2_max when scaled by total body mass and across these test endpoints.

| Test endpoint | RMSE | Pearson's *r* | Spearman's $\rho$ | Pooled bias | Female bias | Male bias |
| --- | --- | --- | --- | --- | --- | --- |
| Stage IV | 6.2 | 0.79 | 0.77 | -3.1 ± 5.5* | -2.9 ± 5.1* | -3.3 ± 5.9* |
| Stage III | 7.3 | 0.67 | 0.64 | -2.5 ± 6.9* | -3.6 ± 6.5* | -1.3 ± 7.2 |
| Stage II | 16.9 | 0.11 | 0.17 | -14.2 ± 9.2* | -13.4 ± 7.6* | -15.2 ± 10.8* |
| 100% age-predicted HRmax | 6.0 | 0.79 | 0.76 | -2.6 ± 5.5* | -2.9 ± 5.7* | -2.2 ± 5.3* |
| 90% age-predicted HRmax | 6.5 | 0.76 | 0.72 | -3.0 ± 5.8* | -3.4 ± 6.1* | -2.6 ± 5.5* |
| 80% age-predicted HRmax | 6.8 | 0.72 | 0.67 | -2.9 ± 6.2* | -3.5 ± 6.3* | -2.3 ± 6.1* |
| 70% age-predicted HRmax | 7.4 | 0.64 | 0.63 | -2.3 ± 7.0* | -3.2 ± 6.8* | -1.5 ± 7.3 |
| 60% age-predicted HRmax | 8.1 | 0.58 | 0.58 | -1.6 ± 8.0 | -3.0 ± 8.0* | -0.1 ± 7.9 |
| 50% age-predicted HRmax | 10.0 | 0.57 | 0.55 | -3.0 ± 9.6* | -6.0 ± 9.4* | 0.1 ± 8.9 |

Age-predicted HRmax = $208 - 0.7 \cdot \text{age}(y)$ ; RMSE: root mean squared error; Bias values are mean ± standard deviation ml O<sub>2</sub>·kg<sup>-1</sup>·min<sup>-1</sup>; \*: One-sample t-test $p < 0.05$

The VO_2_max estimation method developed for the CRTT demonstrated good levels of agreement with measured VO_2_max when the end of stages III and IV were used as test endpoints (Pearson’s *r* range: 0.67 to 0.79; mean bias range: -3.1 to -2.5 ml O_2_·kg^-1^·min^-1^) or when the attainment of 80% to 100% of the participants age-predicted HRmax was used as a test endpoint (Pearson’s *r* range: 0.72 to 0.79; mean bias range: -3.0 to -2.6 ml O_2_·kg^-1^·min^-1^). All other test endpoint criteria examined resulted in worse levels of agreement.

We also examined agreement for estimated VO_2_max when scaled by fat free mass:

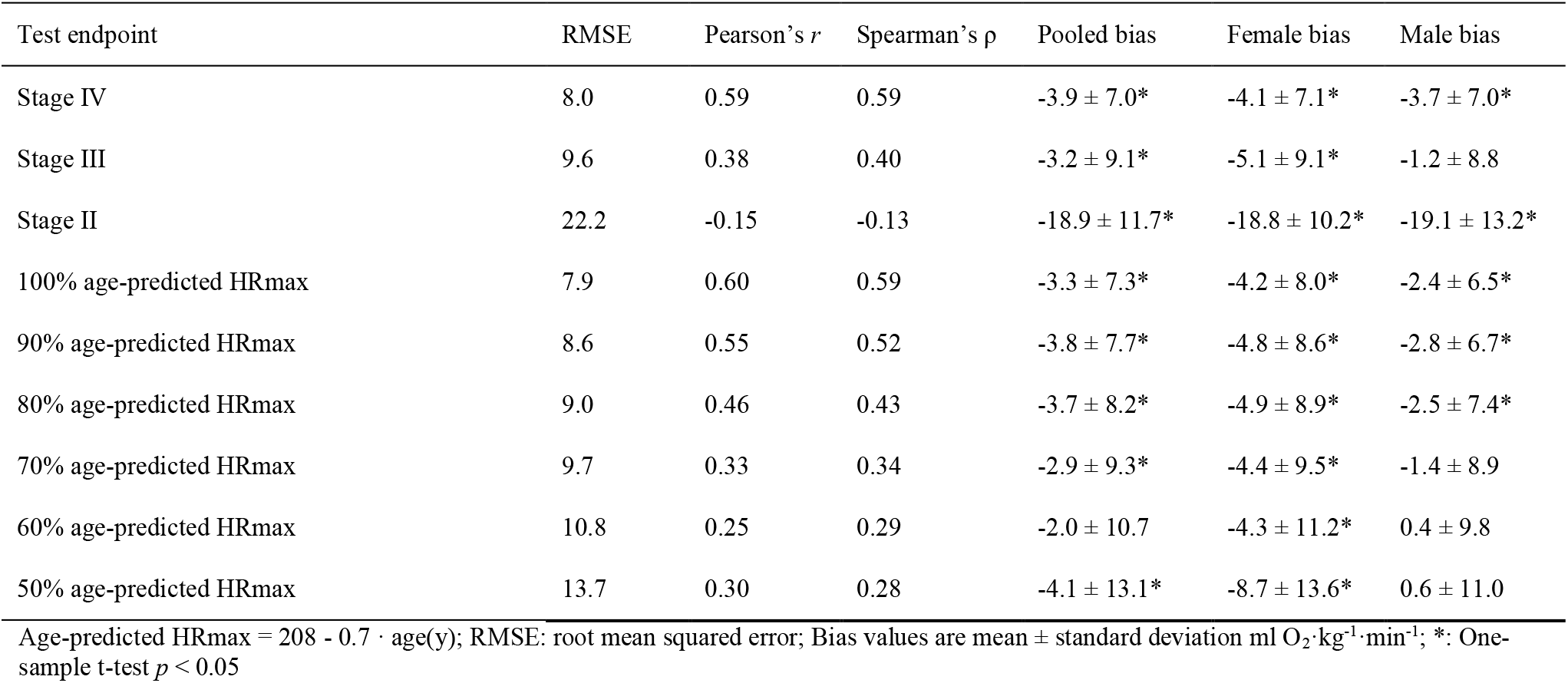

| Test endpoint | RMSE | Pearson's $r$ | Spearman's $\rho$ | Pooled bias | Female bias | Male bias |
| --- | --- | --- | --- | --- | --- | --- |
| Stage IV | 8.0 | 0.59 | 0.59 | $-3.9 \pm 7.0^*$ | $-4.1 \pm 7.1^*$ | $-3.7 \pm 7.0^*$ |
| Stage III | 9.6 | 0.38 | 0.40 | $-3.2 \pm 9.1^*$ | $-5.1 \pm 9.1^*$ | $-1.2 \pm 8.8$ |
| Stage II | 22.2 | -0.15 | -0.13 | $-18.9 \pm 11.7^*$ | $-18.8 \pm 10.2^*$ | $-19.1 \pm 13.2^*$ |
| 100% age-predicted HRmax | 7.9 | 0.60 | 0.59 | $-3.3 \pm 7.3^*$ | $-4.2 \pm 8.0^*$ | $-2.4 \pm 6.5^*$ |
| 90% age-predicted HRmax | 8.6 | 0.55 | 0.52 | $-3.8 \pm 7.7^*$ | $-4.8 \pm 8.6^*$ | $-2.8 \pm 6.7^*$ |
| 80% age-predicted HRmax | 9.0 | 0.46 | 0.43 | $-3.7 \pm 8.2^*$ | $-4.9 \pm 8.9^*$ | $-2.5 \pm 7.4^*$ |
| 70% age-predicted HRmax | 9.7 | 0.33 | 0.34 | $-2.9 \pm 9.3^*$ | $-4.4 \pm 9.5^*$ | $-1.4 \pm 8.9$ |
| 60% age-predicted HRmax | 10.8 | 0.25 | 0.29 | $-2.0 \pm 10.7$ | $-4.3 \pm 11.2^*$ | $0.4 \pm 9.8$ |
| 50% age-predicted HRmax | 13.7 | 0.30 | 0.28 | $-4.1 \pm 13.1^*$ | $-8.7 \pm 13.6^*$ | $0.6 \pm 11.0$ |

Age-predicted HRmax = $208 - 0.7 \cdot \text{age}(\text{y})$ ; RMSE: root mean squared error; Bias values are mean ± standard deviation ml O<sub>2</sub>·kg<sup>-1</sup>·min<sup>-1</sup>; \*: One-sample t-test $p < 0.05$

Levels in agreement were largely similar to those attained when VO_2_max was scaled by total body mass.

In the main study sample, we examined on average which test endpoint criteria were used across age half-decade groups:

| Sex | Women |  |  |  |  |  |  |
| --- | --- | --- | --- | --- | --- | --- | --- |
| Age group (y) | 29-34 | 35-39 | 40-44 | 45-49 | 50-54 | 55-59 | 60-64 |
| N | 184 | 736 | 1210 | 1322 | 1269 | 972 | 283 |
| Age-predicted HRmax | 185 ± 1 | 182 ± 1 | 178 ± 1 | 175 ± 1 | 171 ± 1 | 168 ± 1 | 165 ± 1 |
| HRmax reached | 160 ± 11 | 156 ± 12 | 152 ± 12 | 150 ± 12 | 146 ± 12 | 143 ± 12 | 139 ± 11 |
| % age-predicted HRmax reached | 86 ± 6 | 86 ± 7 | 86 ± 7 | 86 ± 7 | 85 ± 7 | 85 ± 7 | 84 ± 6 |
| Test duration (minutes) | 15 ± 1 | 15 ± 2 | 15 ± 2 | 15 ± 2 | 15 ± 2 | 14 ± 2 | 14 ± 2 |
| Sex | Men |  |  |  |  |  |  |
| Age group (y) | 29-34 | 35-39 | 40-44 | 45-49 | 50-54 | 55-59 | 60-64 |
| N | 161 | 735 | 1010 | 1135 | 1072 | 929 | 274 |
| Age-predicted HRmax | 185 ± 1 | 182 ± 1 | 178 ± 1 | 175 ± 1 | 171 ± 1 | 168 ± 1 | 165 ± 1 |
| HRmax reached | 156 ± 11 | 153 ± 12 | 149 ± 12 | 147 ± 13 | 143 ± 12 | 139 ± 13 | 137 ± 11 |
| % age-predicted HRmax reached | 84 ± 6 | 84 ± 7 | 84 ± 7 | 84 ± 7 | 83 ± 7 | 83 ± 8 | 83 ± 7 |
| Test duration (minutes) | 17 ± 2 | 17 ± 2 | 17 ± 2 | 16 ± 2 | 16 ± 2 | 16 ± 2 | 15 ± 2 |

These results indicate that test endpoint criteria used in the main study participant sample resulted in fitness estimates with acceptable levels of agreement, irrespective of participant age.

**Supplementary Table 1.** Descriptive statistics in women by sociodemographic characteristics. The Fenland Study 2005-2015.

|  | Count (%) | Age | Total body mass (kg) | BMI (kg·m<sup>-2</sup>) | FFM (kg) | RHR (bpm) | VO<sub>2</sub>max<sub>tbm</sub> | VO<sub>2</sub>max<sub>fmm</sub> | PAEE |
| --- | --- | --- | --- | --- | --- | --- | --- | --- | --- |
| Age group (y) |  |  |  |  |  |  |  |  |  |
| 29-34 | 191 (2.9%) | 33 ± 1 | 66.8 ± 12.4 | 24.6 ± 4.2 | 40.8 ± 4.5 | 64 ± 8 | 36.1 ± 6.2 | 58.5 ± 8.8 | 58 ± 21 |
| 35-39 | 770 (11.5%) | 38 ± 1 | 70.2 ± 15.7 | 25.8 ± 5.5 | 42.0 ± 5.6 | 65 ± 9 | 36.2 ± 7.1 | 59.2 ± 10.2 | 57 ± 22 |
| 40-44 | 1281 (19.1%) | 43 ± 1 | 71.7 ± 15.7 | 26.4 ± 5.5 | 42.2 ± 5.7 | 65 ± 9 | 35.8 ± 7.2 | 59.3 ± 10.5 | 52 ± 20 |
| 45-49 | 1433 (21.4%) | 48 ± 1 | 71.5 ± 14.5 | 26.5 ± 5.3 | 42.0 ± 5.3 | 65 ± 9 | 35.5 ± 7.7 | 59.5 ± 11.7 | 50 ± 20 |
| 50-54 | 1451 (21.7%) | 52 ± 1 | 71.8 ± 14.1 | 26.9 ± 5.2 | 41.1 ± 5.1 | 65 ± 9 | 35.1 ± 7.8 | 60.0 ± 12.6 | 47 ± 19 |
| 55-59 | 1182 (17.7%) | 57 ± 1 | 71.5 ± 14.3 | 26.9 ± 5.1 | 40.4 ± 5.2 | 64 ± 9 | 34.9 ± 8.1 | 60.3 ± 13.3 | 44 ± 18 |
| 60-64 | 383 (5.7%) | 61 ± 1 | 71.7 ± 16.1 | 27.2 ± 5.6 | 39.9 ± 5.2 | 64 ± 9 | 34.7 ± 8.4 | 60.0 ± 14.0 | 41 ± 17 |
| Ethnicity |  |  |  |  |  |  |  |  |  |
| White | 6172 (92.2%) | 49 ± 7 | 71.6 ± 14.8 | 26.6 ± 5.3 | 41.5 ± 5.3 | 65 ± 9 | 35.6 ± 7.6 | 59.9 ± 11.8 | 49 ± 20 |
| South Asian | 78 (1.2%) | 46 ± 8 | 62.7 ± 11.4 | 25.4 ± 4.4 | 35.7 ± 4.7 | 68 ± 9 | 34.6 ± 10.4 | 60.2 ± 15.4 | 44 ± 20 |
| Black | 30 (0.4%) | 44 ± 7 | 77.0 ± 19.5 | 28.7 ± 6.6 | 45.6 ± 8.2 | 64 ± 9 | 35.0 ± 8.1 | 56.7 ± 13.1 | 49 ± 22 |
| East Asian | 48 (0.7%) | 47 ± 7 | 58.7 ± 8.5 | 23.3 ± 2.9 | 36.2 ± 4.2 | 66 ± 10 | 38.6 ± 10.3 | 61.7 ± 15.5 | 46 ± 20 |
| Others or unknown | 363 (5.4%) | 44 ± 8 | 70.6 ± 15.2 | 26.3 ± 5.3 | 40.7 ± 5.1 | 64 ± 8 | 33.0 ± 6.8 | 55.0 ± 10.3 | 48 ± 19 |
| Education level |  |  |  |  |  |  |  |  |  |
| None | 48 (0.7%) | 51 ± 6 | 75.5 ± 16.0 | 28.6 ± 6.1 | 42.3 ± 5.6 | 64 ± 9 | 34.6 ± 8.6 | 60.0 ± 15.4 | 46 ± 21 |
| Basic | 1459 (21.8%) | 50 ± 7 | 73.4 ± 15.6 | 27.7 ± 5.6 | 41.3 ± 5.4 | 66 ± 9 | 35.2 ± 8.2 | 60.7 ± 13.2 | 48 ± 21 |
| Further | 3031 (45.3%) | 49 ± 7 | 72.1 ± 15.0 | 26.9 ± 5.4 | 41.4 ± 5.4 | 65 ± 9 | 35.0 ± 7.5 | 59.5 ± 11.8 | 49 ± 20 |
| Higher | 2153 (32.2%) | 48 ± 8 | 68.8 ± 13.6 | 25.2 ± 4.8 | 41.4 ± 5.3 | 64 ± 8 | 36.2 ± 7.3 | 59.2 ± 10.9 | 51 ± 20 |
| Work type |  |  |  |  |  |  |  |  |  |
| Sedentary | 3124 (46.7%) | 48 ± 7 | 71.3 ± 14.8 | 26.4 ± 5.3 | 41.5 ± 5.4 | 65 ± 9 | 35.1 ± 7.2 | 59.1 ± 11.1 | 46 ± 18 |
| Standing | 2093 (31.3%) | 49 ± 7 | 70.8 ± 14.1 | 26.4 ± 5.1 | 41.2 ± 5.3 | 64 ± 9 | 35.7 ± 7.8 | 60.1 ± 12.2 | 52 ± 21 |
| Manual | 501 (7.5%) | 48 ± 7 | 71.2 ± 14.6 | 26.7 ± 5.3 | 41.8 ± 5.3 | 64 ± 9 | 36.4 ± 8.3 | 61.2 ± 13.7 | 59 ± 23 |
| Retired | 240 (3.6%) | 59 ± 4 | 72.0 ± 14.9 | 27.4 ± 5.3 | 40.0 ± 5.2 | 66 ± 9 | 34.5 ± 7.8 | 60.2 ± 13.8 | 42 ± 17 |
| Unemployed | 77 (1.2%) | 47 ± 7 | 74.9 ± 18.4 | 27.9 ± 6.5 | 42.1 ± 6.6 | 66 ± 9 | 35.3 ± 8.3 | 61.1 ± 13.1 | 45 ± 19 |
| Unknown | 656 (9.8%) | 47 ± 8 | 72.6 ± 16.8 | 27.0 ± 6.1 | 41.6 ± 5.3 | 66 ± 9 | 35.5 ± 8.4 | 59.2 ± 12.1 | 51 ± 22 |
| Income |  |  |  |  |  |  |  |  |  |
| <£20 000 | 1103 (16.5%) | 49 ± 8 | 72.9 ± 15.9 | 27.5 ± 5.7 | 41.2 ± 5.5 | 66 ± 9 | 35.4 ± 8.0 | 60.6 ± 12.8 | 47 ± 21 |
| £20 000 - £40 000 | 2367 (35.4%) | 49 ± 8 | 71.8 ± 15.1 | 26.8 ± 5.5 | 41.3 ± 5.4 | 65 ± 9 | 35.2 ± 7.6 | 59.8 ± 12.0 | 49 ± 20 |
| >£40 000 | 2995 (44.8%) | 48 ± 7 | 70.3 ± 14.0 | 25.9 ± 5.0 | 41.5 ± 5.2 | 64 ± 9 | 35.7 ± 7.2 | 59.2 ± 11.0 | 51 ± 19 |
| Unknown | 226 (3.4%) | 51 ± 7 | 72.5 ± 16.2 | 27.6 ± 5.8 | 40.8 ± 5.7 | 66 ± 10 | 35.8 ± 10.3 | 61.1 ± 16.6 | 48 ± 23 |
| Marital status |  |  |  |  |  |  |  |  |  |
| Single | 438 (6.6%) | 46 ± 7 | 72.4 ± 16.3 | 26.6 ± 5.8 | 41.9 ± 5.8 | 65 ± 9 | 36.3 ± 7.3 | 60.8 ± 11.9 | 51 ± 21 |
| Married/living as married | 4114 (61.6%) | 50 ± 7 | 71.0 ± 14.3 | 26.4 ± 5.2 | 41.3 ± 5.3 | 65 ± 9 | 35.7 ± 7.6 | 60.0 ± 11.9 | 49 ± 20 |
| Widowed/separated/divorced | 593 (8.9%) | 51 ± 7 | 71.5 ± 15.2 | 26.7 ± 5.4 | 41.2 ± 5.3 | 65 ± 9 | 36.2 ± 7.9 | 61.1 ± 12.4 | 48 ± 20 |
| Unknown | 1538 (23.0%) | 46 ± 7 | 71.9 ± 15.7 | 26.8 ± 5.6 | 41.6 ± 5.5 | 64 ± 8 | 34.2 ± 7.4 | 57.6 ± 11.2 | 50 ± 21 |
| Smoker status |  |  |  |  |  |  |  |  |  |
| Never smoked | 3737 (55.9%) | 49 ± 7 | 70.5 ± 14.7 | 26.3 ± 5.3 | 41.0 ± 5.4 | 65 ± 9 | 35.0 ± 7.4 | 58.9 ± 11.5 | 48 ± 19 |
| Ex-smoker | 2141 (32.0%) | 49 ± 8 | 72.7 ± 15.0 | 27.0 ± 5.4 | 41.9 ± 5.3 | 64 ± 8 | 35.3 ± 7.4 | 59.7 ± 11.7 | 49 ± 20 |
| Current smoker | 727 (10.9%) | 47 ± 7 | 71.3 ± 14.6 | 26.6 ± 5.3 | 41.6 ± 5.4 | 64 ± 9 | 38.1 ± 8.5 | 63.7 ± 13.3 | 56 ± 24 |
| Unknown | 78 (1.2%) | 48 ± 7 | 71.8 ± 13.6 | 26.6 ± 5.3 | 42.0 ± 4.9 | 62 ± 8 | 34.7 ± 6.2 | 58.5 ± 10.8 | 52 ± 23 |
| Test site |  |  |  |  |  |  |  |  |  |
| Cambridge | 2329 (34.8%) | 49 ± 7 | 69.3 ± 13.5 | 25.5 ± 4.8 | 40.9 ± 5.3 | 64 ± 9 | 35.8 ± 7.7 | 59.5 ± 11.7 | 50 ± 20 |
| Ely | 2557 (38.2%) | 48 ± 7 | 71.7 ± 14.9 | 26.7 ± 5.4 | 41.9 ± 5.3 | 65 ± 9 | 35.3 ± 7.5 | 59.1 ± 11.9 | 49 ± 20 |
| Wisbech | 1805 (27.0%) | 49 ± 7 | 73.5 ± 15.9 | 27.6 ± 5.6 | 41.4 ± 5.5 | 66 ± 9 | 35.1 ± 7.6 | 60.8 ± 11.9 | 49 ± 20 |
| BMI group (kg·m<sup>-2</sup>) |  |  |  |  |  |  |  |  |  |
| <25 | 3085 (46.1%) | 48 ± 8 | 60.7 ± 6.3 | 22.3 ± 1.7 | 38.8 ± 4.1 | 64 ± 8 | 37.5 ± 7.5 | 58.3 ± 11.0 | 55 ± 21 |
| 25-30 | 2159 (32.3%) | 49 ± 7 | 72.6 ± 6.7 | 27.1 ± 1.4 | 41.5 ± 4.4 | 65 ± 9 | 34.8 ± 7.2 | 60.7 ± 12.3 | 48 ± 18 |
| >30 | 1444 (21.6%) | 50 ± 7 | 92.2 ± 13.6 | 34.6 ± 4.5 | 46.8 ± 5.2 | 67 ± 9 | 31.5 ± 6.9 | 61.1 ± 12.8 | 39 ± 16 |
| PAEE group (kJ·day<sup>-1</sup>·kg<sup>-1</sup>) |  |  |  |  |  |  |  |  |  |
| <40 | 2341 (35.0%) | 50 ± 7 | 76.2 ± 17.3 | 28.5 ± 6.2 | 41.8 ± 6.0 | 68 ± 9 | 32.4 ± 7.4 | 57.5 ± 12.7 | 30 ± 7 |
| 40-60 | 2529 (37.8%) | 48 ± 7 | 70.2 ± 13.4 | 26.1 ± 4.7 | 41.0 ± 5.1 | 64 ± 8 | 35.4 ± 6.9 | 59.6 ± 11.3 | 49 ± 6 |
| >60 | 1821 (27.2%) | 47 ± 7 | 66.7 ± 11.1 | 24.5 ± 3.9 | 41.4 ± 4.8 | 61 ± 8 | 39.0 ± 7.2 | 62.1 ± 11.0 | 75 ± 14 |

Values are mean ± standard deviation, unless otherwise indicated. BMI: Body mass index. FFM: Fat free mass. SBP: Systolic blood pressure. DBP: Diastolic blood pressure. RHR: Resting heart rate. VO<sub>2</sub>max<sub>tbm</sub>: Maximal oxygen consumption per kilogram total body mass (ml O<sub>2</sub>·min<sup>-1</sup>·kg<sup>-1</sup>). VO<sub>2</sub>max<sub>fmm</sub>: Maximal oxygen consumption per kilogram fat free mass (ml O<sub>2</sub>·min<sup>-1</sup>·kg<sup>-1</sup>). PAEE: Physical activity energy expenditure (kJ·day<sup>-1</sup>·kg<sup>-1</sup>).

**Supplementary Table 2.**
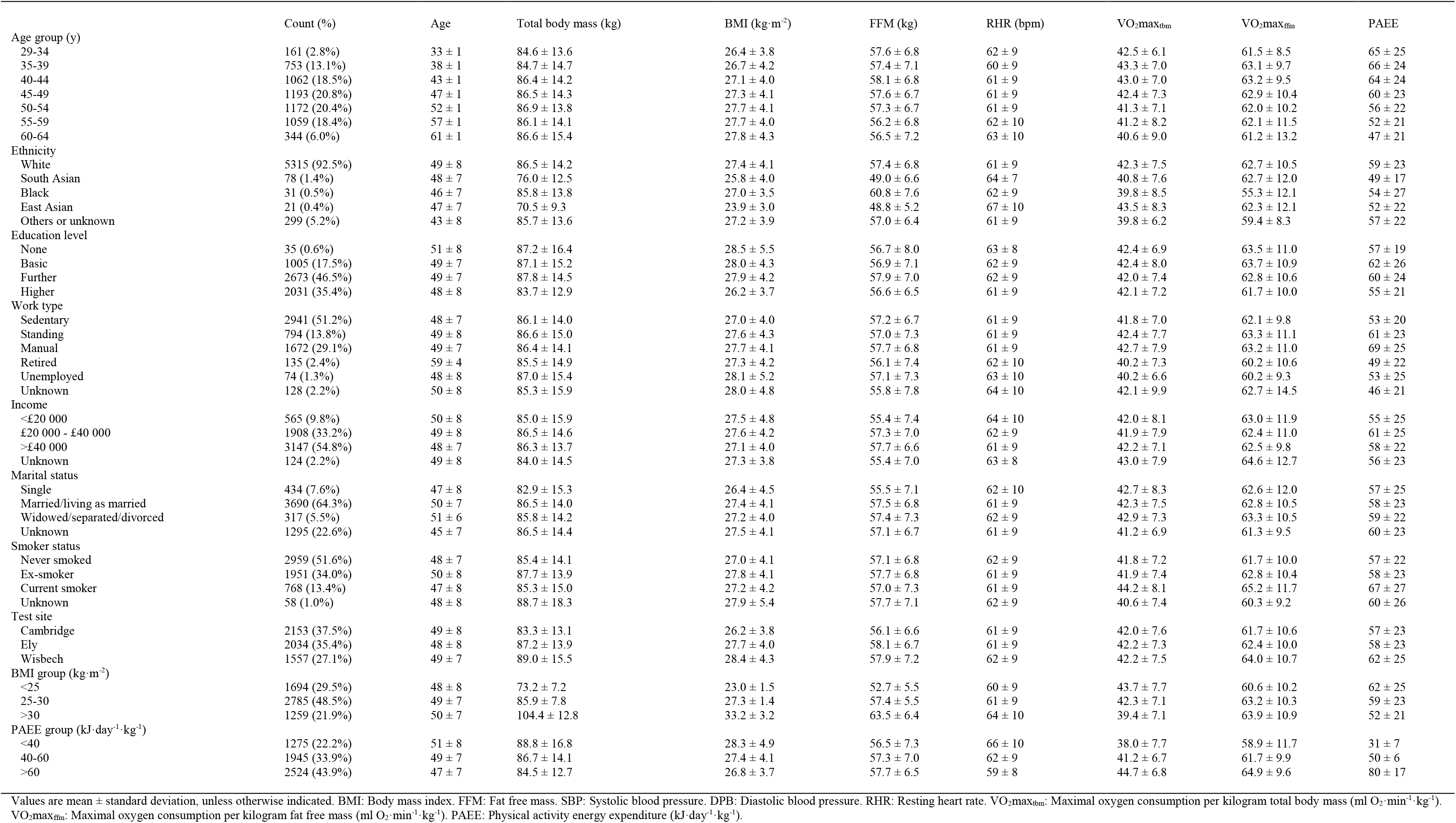
Descriptive statistics in men by sociodemographic characteristics. The Fenland Study 2005-2015.

**Supplemental Figure 1.**
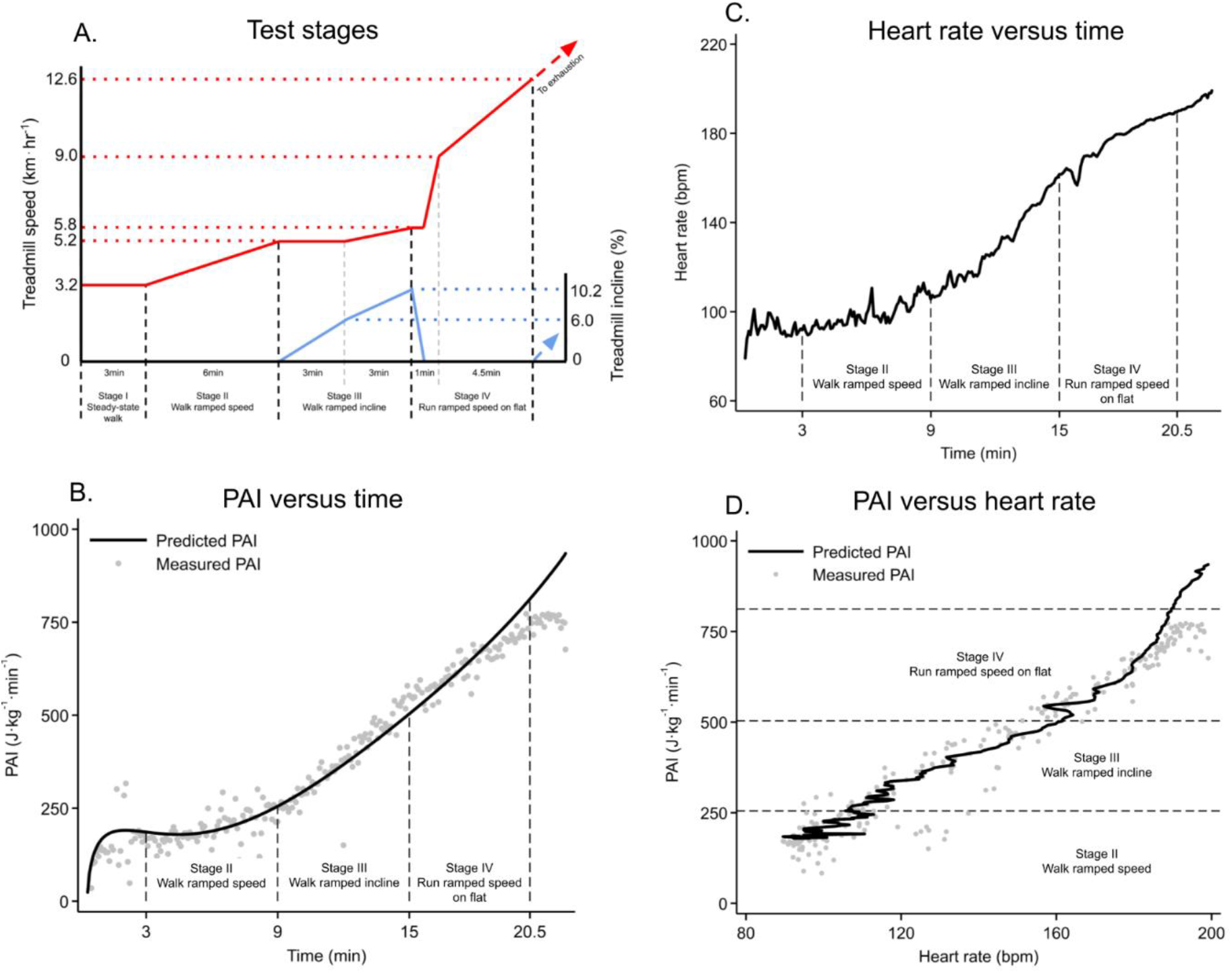
A: Treadmill speed (solid red lines) and incline (solid blue lines) over different stages (time periods between dashed black vertical lines) of the Cambridge Ramped Treadmill Test (CRTT). The test has four stages. The first stage consists of walking at 3.2 km·h^-1^ at 0% incline for 3 min. The second stage consists of walking while treadmill speed increases from 3.2 to 5.2 km·h^-1^ at 0% incline for 6 min. The third stage consists of walking at 5.2 km·h^-1^ while incline increases from 0 to 6% for 3 min, and then walking while treadmill speed increases from 5.2 to 5.8 km·h^-1^ and incline from 6 to 10.2% for 3 min. The fourth consists of running while treadmill speed increases from 5.8 to 9.0 km·h^-1^ and incline decreases to 0% for 1 min, and then running while treadmill speed increases from 9.0 to 12.6 km·h^-1^ and at 0% incline for 4.5 min. Instantaneous work rate values, expressed as physical activity intensity (J·kg^-1^·min^-1^), were computed from treadmill speed and incline according to measured physical activity intensity. Dotted red horizontal lines represent treadmill speeds. Dotted blue horizontal lines represent treadmill inclines. Dashed vertical grey lines divide sub-phases within stages III and IV. To measure VO_2_max in the validation substudy, stage IV was extended (dashed red and blue arrowed lines) until exhaustion was achieved. B: Exemplar data of predicted (solid line) and directly measured (grey dots) physical activity intensity (PAI) across stages II-IV. C: Exemplar data of heart rate across stages II-IV. D: Exemplar data demonstrating PAI-to-heart rate relationship across stages II-IV.

**Supplemental Figure 2.**
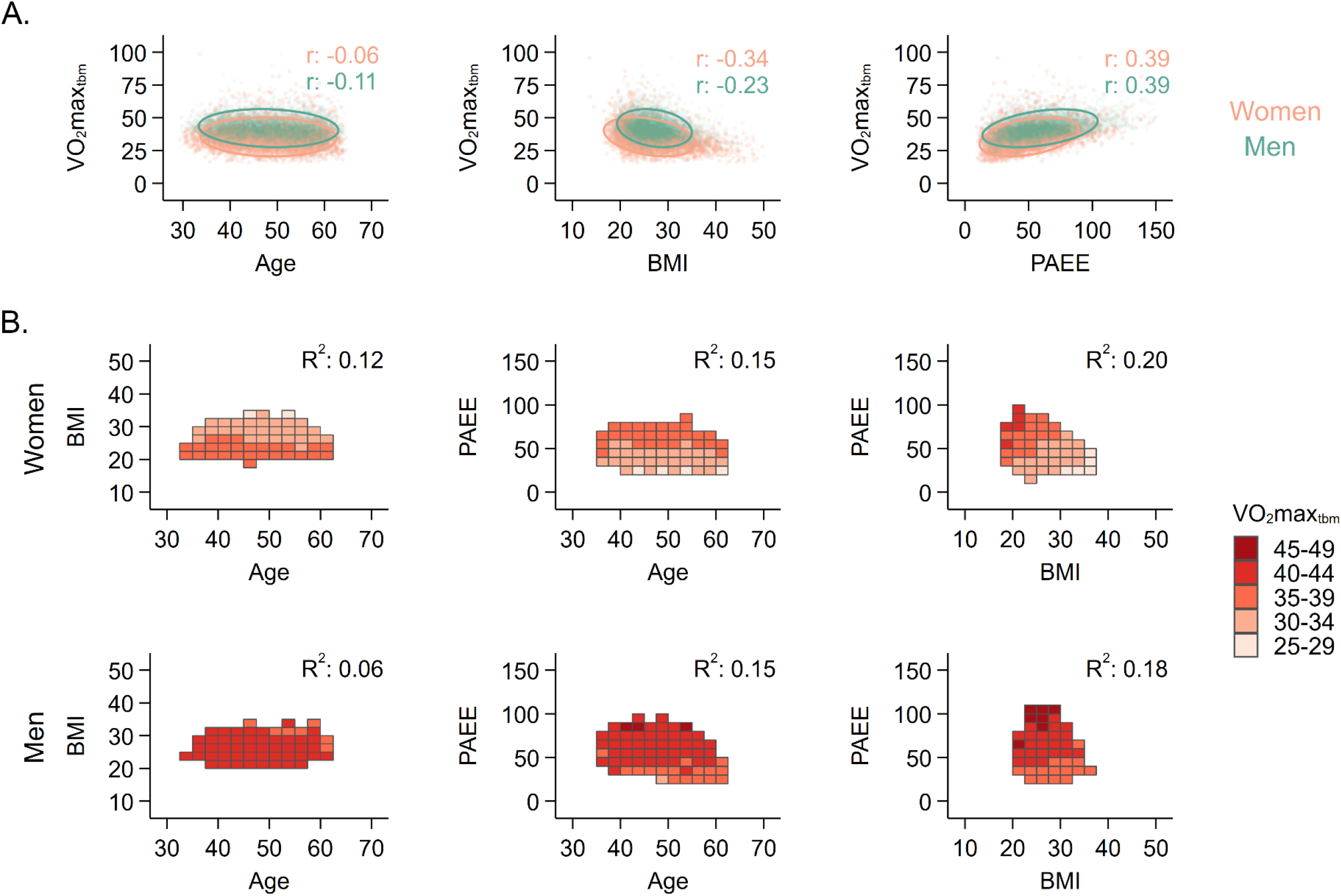
A: Scatterplots and 95% confidence interval ellipses demonstrating univariate associations of VO_2_max_tbm_ with age, BMI, and PAEE. Dots represent values for individual participants. r values are Pearson correlation coefficients. B: Heat plots demonstrating bivariate associations of VO_2_max_tbm_ with age, BMI, and PAEE. Values for rectangular fields were computed as the median VO_2_max_tbm_ for all participants within a given range on each axis. Rectangular fields were omitted if less than 30 participants fell within that range. R^2^ values were computed by multiple linear regression using VO_2_max_tbm_ as the dependent variable.

**Supplemental Figure 3.**
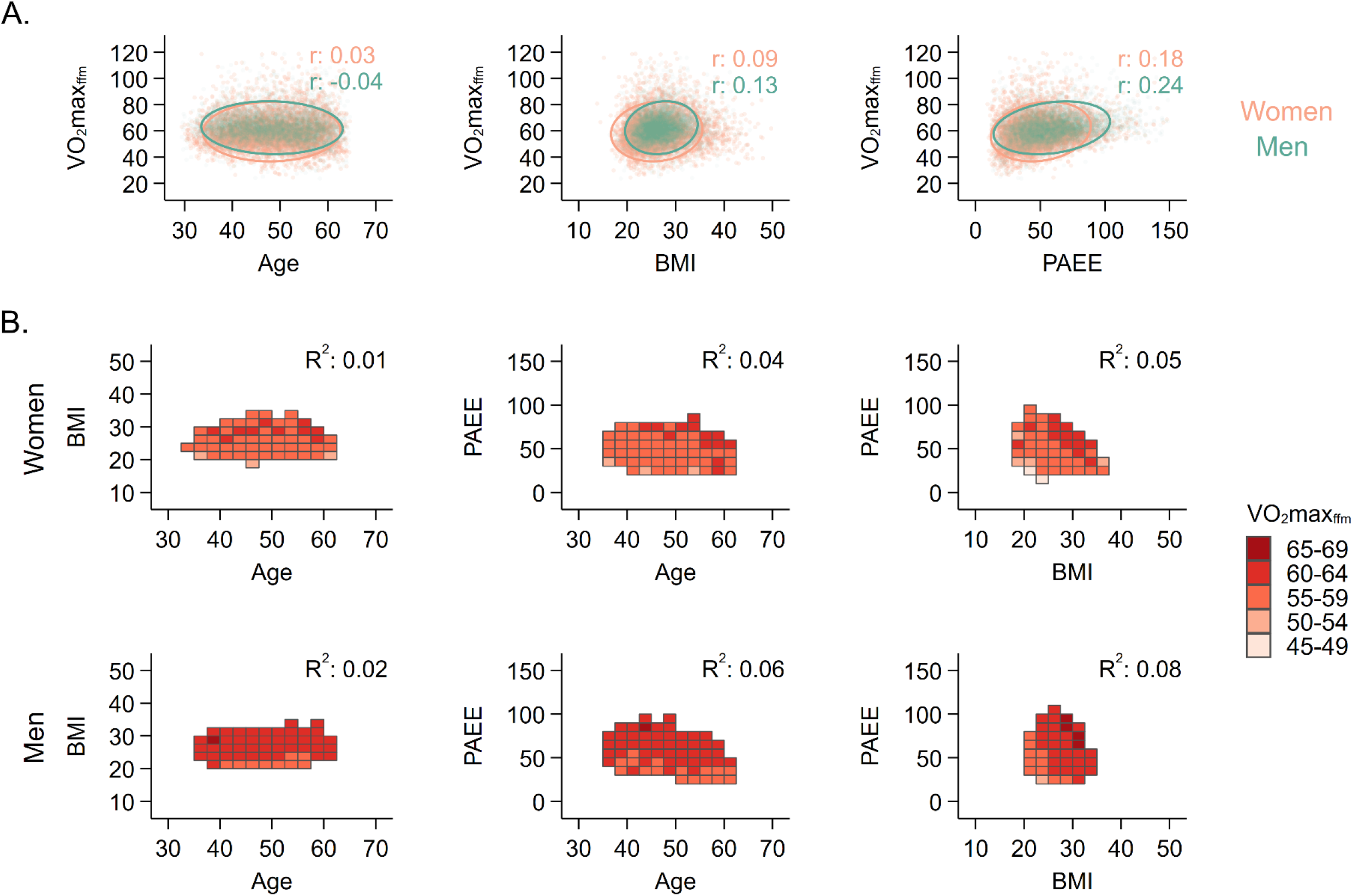
A: Scatterplots and 95% confidence interval ellipses demonstrating univariate associations of VO_2_max_ffm_ with age, BMI, and PAEE. Dots represent values for individual participants. r values are Pearson correlation coefficients. B: Heat plots demonstrating bivariate associations of VO_2_max_ffm_ with age, BMI, and PAEE. Values for rectangular fields were computed as the median VO_2_max_ffm_ for all participants within a given range on each axis. Rectangular fields were omitted if less than 30 participants fell within that range. R^2^ values were computed by multiple linear regression using VO_2_max_ffm_ as the dependent variable.

## Notes

### Competing Interest Statement

The authors have declared no competing interest.

### Author Declarations

The Health Research Authority NRES Committee East of England-Cambridge Central approved the study in accordance with the Declaration of Helsinki. All participants gave written informed consent. The Fenland Study has a dedicated Patient and Public Involvement panel, who provided input on the acceptability of the study protocols and participant data confidentiality. This study complied with the items listed in the Strengthening the Reporting of Observational Studies in Epidemiology (STROBE) guidelines.

